# Spatial Transcriptomic Analysis of Pituitary Corticotroph Tumors Unveils Intratumor Heterogeneity

**DOI:** 10.1101/2023.08.04.23293576

**Authors:** Jeremie Oliver Piña, Fabio R. Faucz, Cameron Padilla, Charalampos S. Floudas, Prashant Chittiboina, Martha Quezado, Christina Tatsi

**Affiliations:** Section on Craniofacial Genetic Disorders, Eunice Kennedy Shriver National Institute of Child Health and Human Development, National Institutes of Health, Bethesda, Maryland, USA; Molecular Genomics Core, Eunice Kennedy Shriver National Institute of Child Health and Human Development, National Institutes of Health, Bethesda, Maryland, USA; Center for Immuno-Oncology, Center for Cancer Research, National Cancer Institute, National Institutes of Health, Bethesda, MD; Neurosurgery Unit for Pituitary and Inheritable Diseases, National Institute of Neurological Disorders and Stroke, National Institutes of Health, Bethesda, Maryland, USA; Laboratory of Pathology, Center for Cancer Research, National Institutes of Health, Bethesda, Maryland, USA; Unit on Hypothalamic and Pituitary Disorders, Eunice Kennedy Shriver National Institute of Child Health and Human Development, National Institutes of Health, Bethesda, Maryland, USA

**Keywords:** Transcriptomics, Cushing disease, Pituitary Neuroendocrine Tumor

## Abstract

Spatial transcriptomic (ST) analysis of tumors provides a novel approach on studying gene expression along with the localization of tumor cells in their environment to uncover spatial interactions. Herein, we present ST analysis of corticotroph pituitary neuroendocrine tumors (PitNETs) from formalin-fixed, paraffin-embedded (FFPE) tissues. We report that the in situ annotation of tumor tissue can be inferred from the gene expression profiles and is in concordance with the annotation made by a pathologist. Furthermore, relative gene expression in the tumor corresponds to common protein staining used in the evaluation of PitNETs, such as reticulin and Ki-67 index. Finally, we identify intratumor heterogeneity; clusters within the same tumor may present with different secretory capacity and transcriptomic profiles, unveiling potential intratumor cell variability with possible therapeutic interest. Together, our results provide the first attempt to clarify the spatial cell profile in PitNETs.

## Introduction

Pituitary neuroendocrine tumors (PitNETs) are intracranial tumors presenting in up to 10-25% of autopsy cases in adults.(1) However, clinically significant PitNETs involving hormone excess or invasion of surrounding structures are rare. For example, the incidence of adrenocorticotropin (ACTH)-secreting (corticotroph) PitNETs leading to Cushing disease (CD) is 1.5-7.6 cases per million people depending on the population studied; less than 10% of these correspond to children.(2)

Analysis of the genetic background of CD has revealed that somatic or less commonly germline gene defects in *USP8, MEN1, TP53, USP48,* and other genes are found in up to half of the patients; in the remaining cases the cause remains unknown.(3–5) In conjunction with DNA analyses, studies of gene expression have also been used to investigate potential tumorigenesis pathways and therapeutic targets. Over the last years, single cell-RNA sequencing (scRNA-seq) of PitNETs has provided insight on their transcriptional profile delineating differences among various cell lineages or based on their characteristics.(6–9) Few studies on intratumor heterogeneity of PitNETs have been reported with conflicting results and the spatial resolution of cell heterogeneity has not been previously described.(8,9)

Spatial transcriptomics (ST) provide a novel and unique method to study tumors by integrating transcriptional profiling with in situ localization of cell clusters with differential expression in relation to the surrounding tissues.(10) This is useful in identifying intratumor heterogeneity and studying the interaction of tumor cells with their microenvironment, such as the lymphatic or vascular structures; these features may contribute to the aggressive potential of the tumor or unveil cell populations of interest.(11,12) ST techniques are also especially useful in the study of small tumors where tumor availability for extensive RNA-seq studies may be limited and biomarkers for disease prognosis are derived from limited immunostaining targets.

We investigated the ST signature of corticotroph PitNETs derived from formalin-fixed, paraffin-embedded (FFPE) tissues. We reported concordance of tumor identification with pathology annotations but also higher level of subclassification of tumor cells with variable expression profiles.

## Results

### Cohort and sample characteristics

All patients presented with CD during childhood or adolescence, with median age of initial diagnosis of 14.2 years and median age at time of surgical intervention of 15.4 years (range 8.5 – 16.7) (Table 1). One patient (2D) had unusual presentation of a corticotroph PitNET extending to the pituitary stalk that required hypophysectomy. She later presented with recurrent disease at the stalk; only the sample from reoperation was available for analysis. One sample from each patient was processed for ST analysis.

**Table 1.**
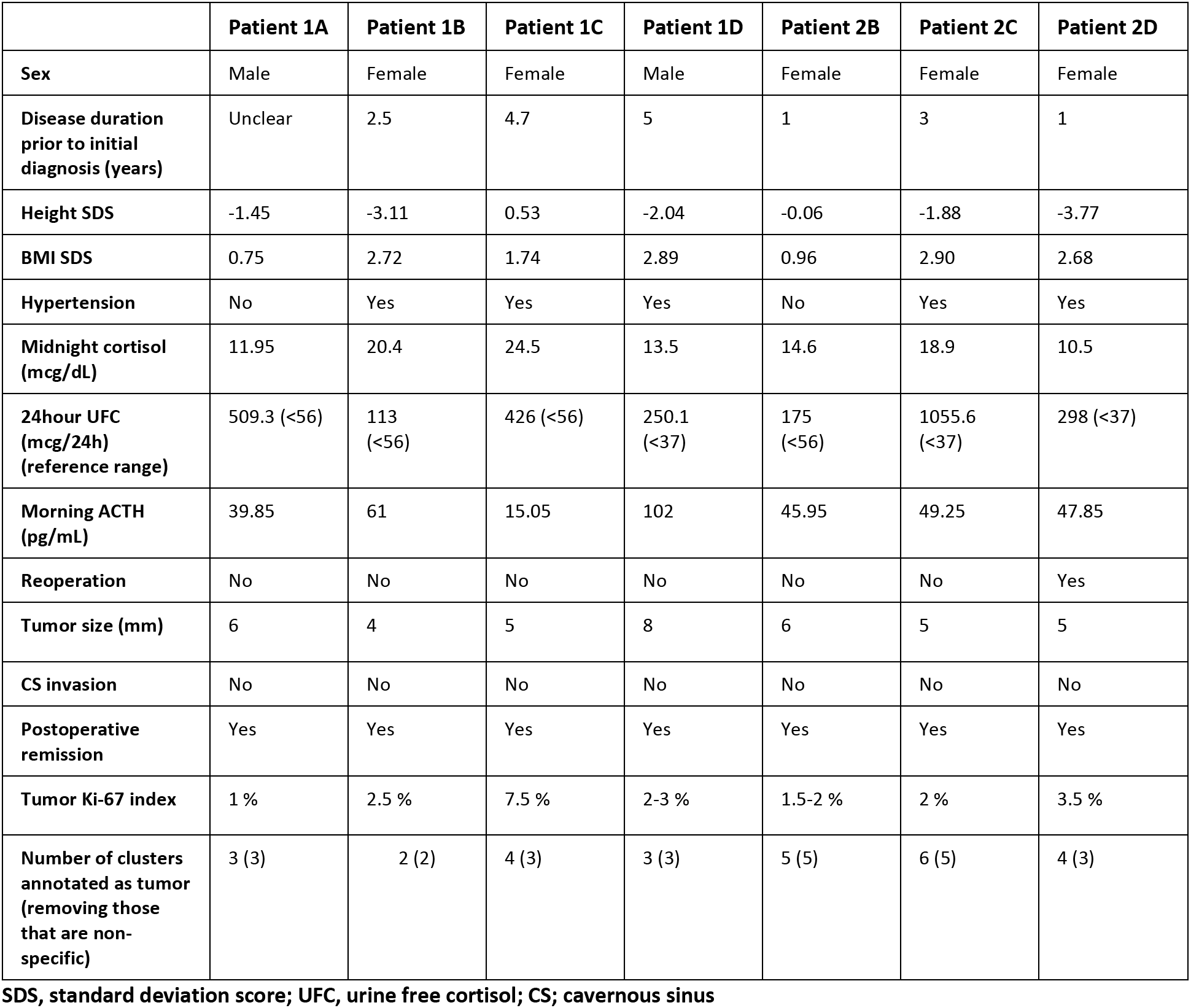
Clinical and biochemical characteristics of patients.

|  | Patient 1A | Patient 1B | Patient 1C | Patient 1D | Patient 2B | Patient 2C | Patient 2D |
| --- | --- | --- | --- | --- | --- | --- | --- |
| Sex | Male | Female | Female | Male | Female | Female | Female |
| Disease duration prior to initial diagnosis (years) | Unclear | 2.5 | 4.7 | 5 | 1 | 3 | 1 |
| Height SDS | -1.45 | -3.11 | 0.53 | -2.04 | -0.06 | -1.88 | -3.77 |
| BMI SDS | 0.75 | 2.72 | 1.74 | 2.89 | 0.96 | 2.90 | 2.68 |
| Hypertension | No | Yes | Yes | Yes | No | Yes | Yes |
| Midnight cortisol (mcg/dL) | 11.95 | 20.4 | 24.5 | 13.5 | 14.6 | 18.9 | 10.5 |
| 24hour UFC (mcg/24h) (reference range) | 509.3 (<56) | 113 (<56) | 426 (<56) | 250.1 (<37) | 175 (<56) | 1055.6 (<37) | 298 (<37) |
| Morning ACTH (pg/mL) | 39.85 | 61 | 15.05 | 102 | 45.95 | 49.25 | 47.85 |
| Reoperation | No | No | No | No | No | No | Yes |
| Tumor size (mm) | 6 | 4 | 5 | 8 | 6 | 5 | 5 |
| CS invasion | No | No | No | No | No | No | No |
| Postoperative remission | Yes | Yes | Yes | Yes | Yes | Yes | Yes |
| Tumor Ki-67 index | 1 % | 2.5 % | 7.5 % | 2-3 % | 1.5-2 % | 2 % | 3.5 % |
| Number of clusters annotated as tumor (removing those that are non-specific) | 3 (3) | 2 (2) | 4 (3) | 3 (3) | 5 (5) | 6 (5) | 4 (3) |

SDS, standard deviation score; UFC, urine free cortisol; CS; cavernous sinus

### Concordance of tissue clustering with pathology annotations

The reported clusters for each ST slide were reviewed and were annotated as “tumor” based on increased relative expression of marker genes of corticotroph cells (*POMC*, *CRHR1*, *TBX19*, *PCSK1*, and/or *AVPR1B*), decreased relative expression of marker genes of other pituitary lineages (such as *PRL*, *GH1*, *DLK1*, and other) and review of the top differentially expressed genes in each cluster. H&E, reticulin and other relevant immunohistochemistry stains (such as ACTH, Prolactin etc.), where available, were reviewed by an expert pituitary pathologist who annotated the tumor area in each slide.

The pathologist’s annotations and the gene expression-based cluster annotation were concordant (Fig. 1, Supplementary Fig. 1). As expected, the tumor clusters had clear upregulation of marker genes of corticotroph cells and downregulation of marker genes of other pituitary lineages; in addition, UMAP plots of the ST spots showed clear separation of tumor and non-tumor clusters (Fig. 1, Supplementary Fig. 1).

**Fig. 1.**
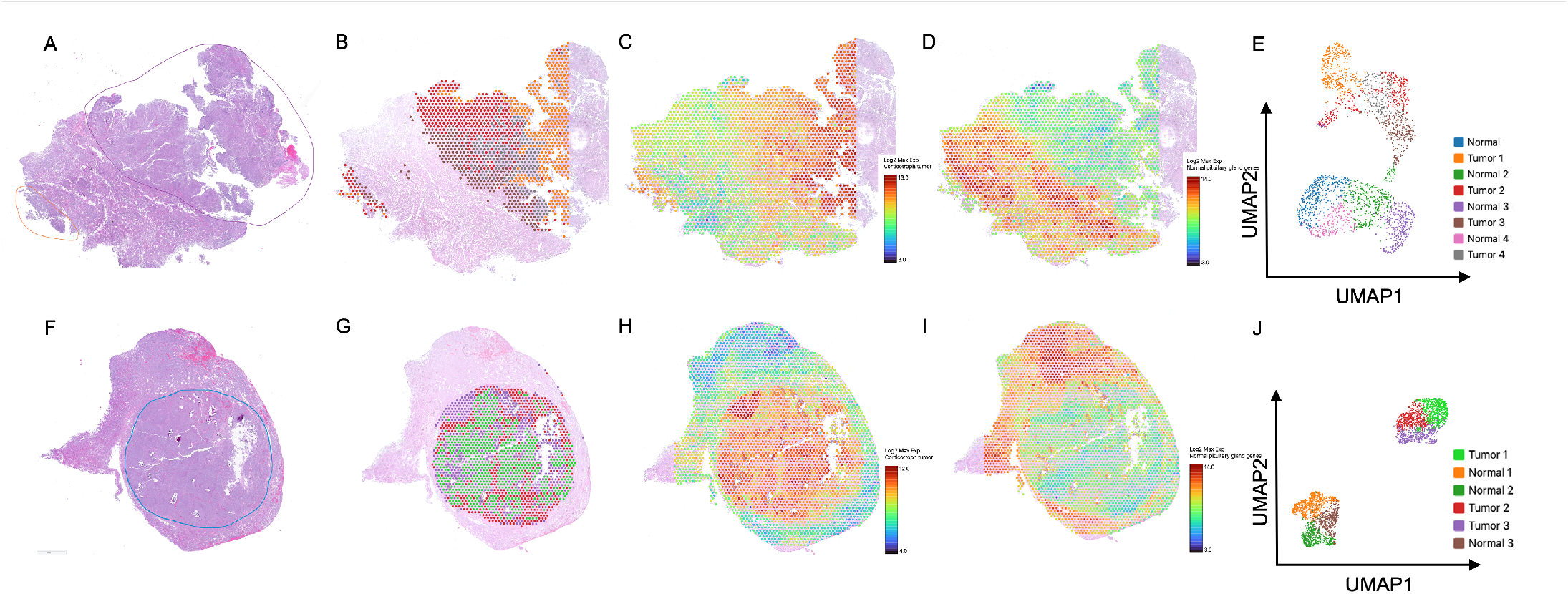
In situ annotation and expression profiles of PitNETs. (A, F) H&E stained slides with tumor annotation made by a pathologist (marked areas). (B, G) In situ cell clusters annotated as tumor (highlighted areas). (C, H) Gene expression of marker genes of corticotroph cells. (D, I) Gene expression of marker genes of non-corticotroph cell lineages of the pituitary gland. (E, J) UMAP graph of clusters annotated as tumor or normal gland.

Because each ST spot may include 1-10 cells, in some cases the spots covering areas of transition from tumor to non-tumor pituitary may be capturing a mixed population of tumor and normal cells. Such areas were hard to annotate based on gene expression profile but were also challenging to accurately annotate in histopathology slides as well. For example, in tumor 1B (Row 2 in Supp. Fig. 1), a group of spots in the periphery shows upregulation of the non-corticotroph pituitary genes without apparent downregulation of the corticotroph marker genes; during clustering most of these spots were incorporated within Tumor cluster 1.

An additional histologic hallmark of PitNETs is the breakdown of reticulin (type III collagen) framework, therefore we reviewed the relative expression profile of reticulin (*COL3A1*) and fibroblast marker genes (*DCN*, *COL1A1*, *LUM*). Expression of these genes was downregulated in tumor areas compared to adjacent normal gland, mirroring the reticulin immunohistochemistry staining pattern, and therefore may also be used in annotation of cell clusters (Fig. 2).

**Fig. 2.**
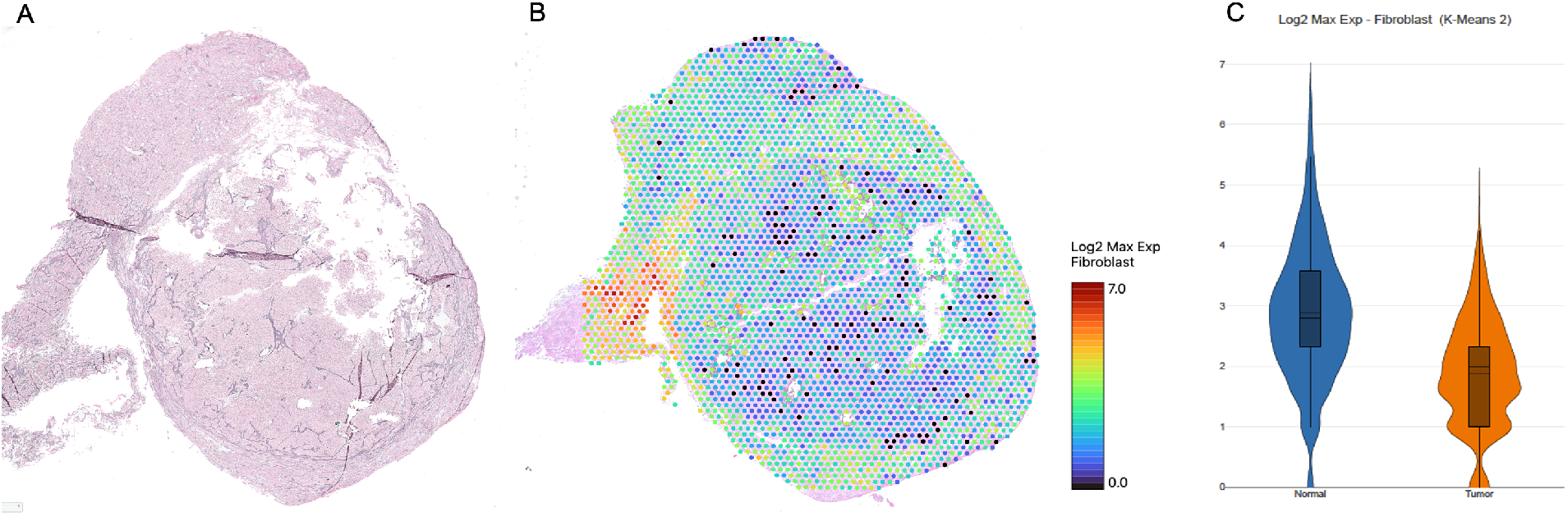
Concordance of protein and gene expression of certain tumor markers. (A) Reticulin staining of a PitNET showing breakdown of reticulin framework inside the tumor area. (B) In situ gene expression of fibroblast marker genes. (C) Violin plot of relative gene expression levels of fibroblast marker genes of tumor versus non-corticotroph cell lineages of the pituitary gland.

### Ki-67 index on spatial transcriptomic analysis

We further investigated if ST analysis can provide information on a marker of aggressive behavior of PitNETs, Ki-67 index. This index is calculated on immunohistochemistry (IHC) staining of the tumors using the MIB1 antibody against Ki-67 protein, and is considered elevated if its value is above >3%, suggesting a more aggressive pituitary tumor.(13) We stained our samples for Ki-67 and identified samples with an elevated Ki-67 index. Review of the relative expression of *MKI67*, the gene encoding the Ki-67 protein, showed that tumors with increased Ki-67 staining also had high expression of *MKI67*, and conversely, tumors with low Ki-67 staining had low *MKI67* expression (Fig. 3), indicating that quantitative *MKI67* mRNA determination may be considered an alternative to IHC-based manual Ki-67 index assessment.

**Fig. 3.**
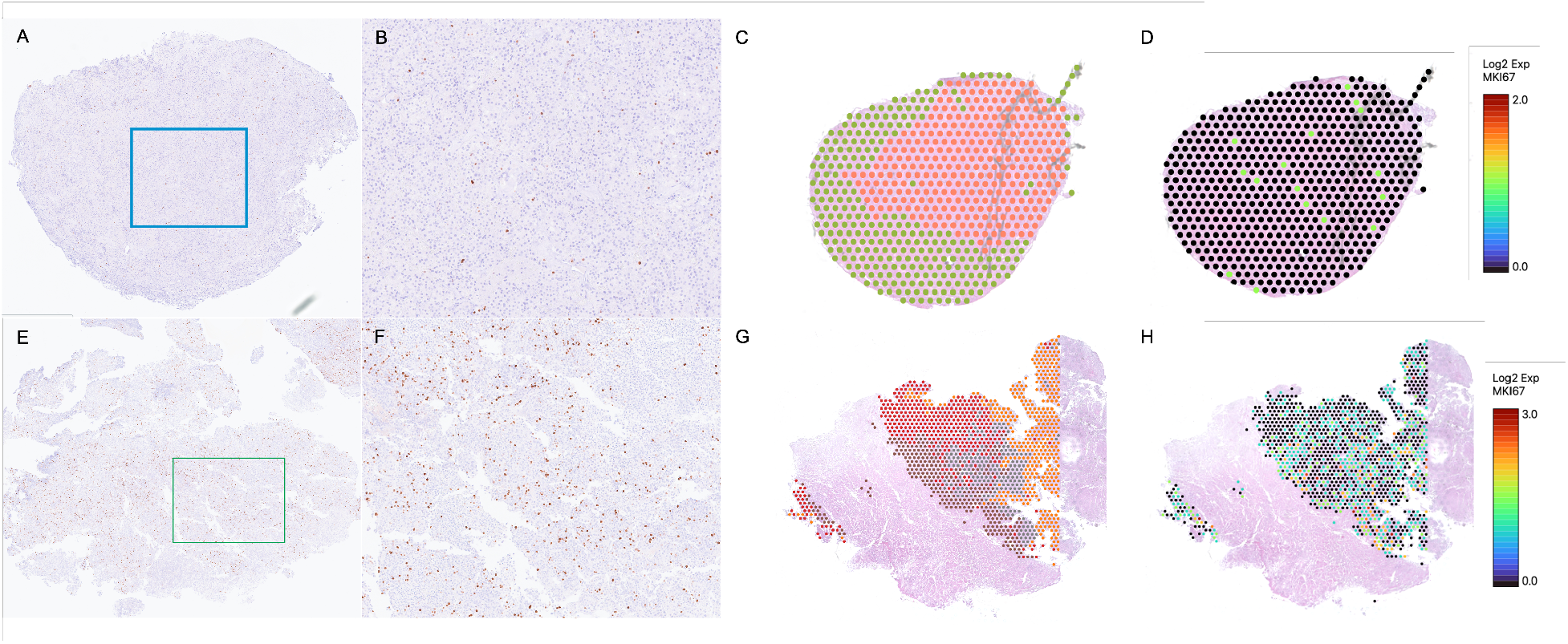
In situ identification of a proliferation marker. (A, B, E, F) MIB1 staining showing a tumor with low staining (A, B) and a tumor with increased staining (E, F). (B) and (F) are magnification of the area within the marked rectangle in figures (A) and (E) respectively. (C, G) In situ cell clusters annotated as tumor (highlighted areas). (D, H) In situ *MKI67* gene expression inside the tumor clusters.

### Intratumor clustering - Cell heterogeneity

Within each tumor area, ST analysis noted a median of 4 tumor clusters (range: 2-6, depicted with different color shown in second columns of Fig. 1 and Supplementary Fig. 1). In three samples, at least one tumor cluster included tissue that appeared damaged probably during resection or sectioning and could represent artifact. When removing these, there were a median of 3 clusters (range: 2-5) per tumor tissue. Some clusters corresponded to specific areas of the tumor, such as orange and red clusters of tumor 2B (Fig. 4C) whereas others represented diffuse cell populations within the tumor tissue (green and purple clusters of tumor 2B, Fig.4C).

**Fig. 4.**
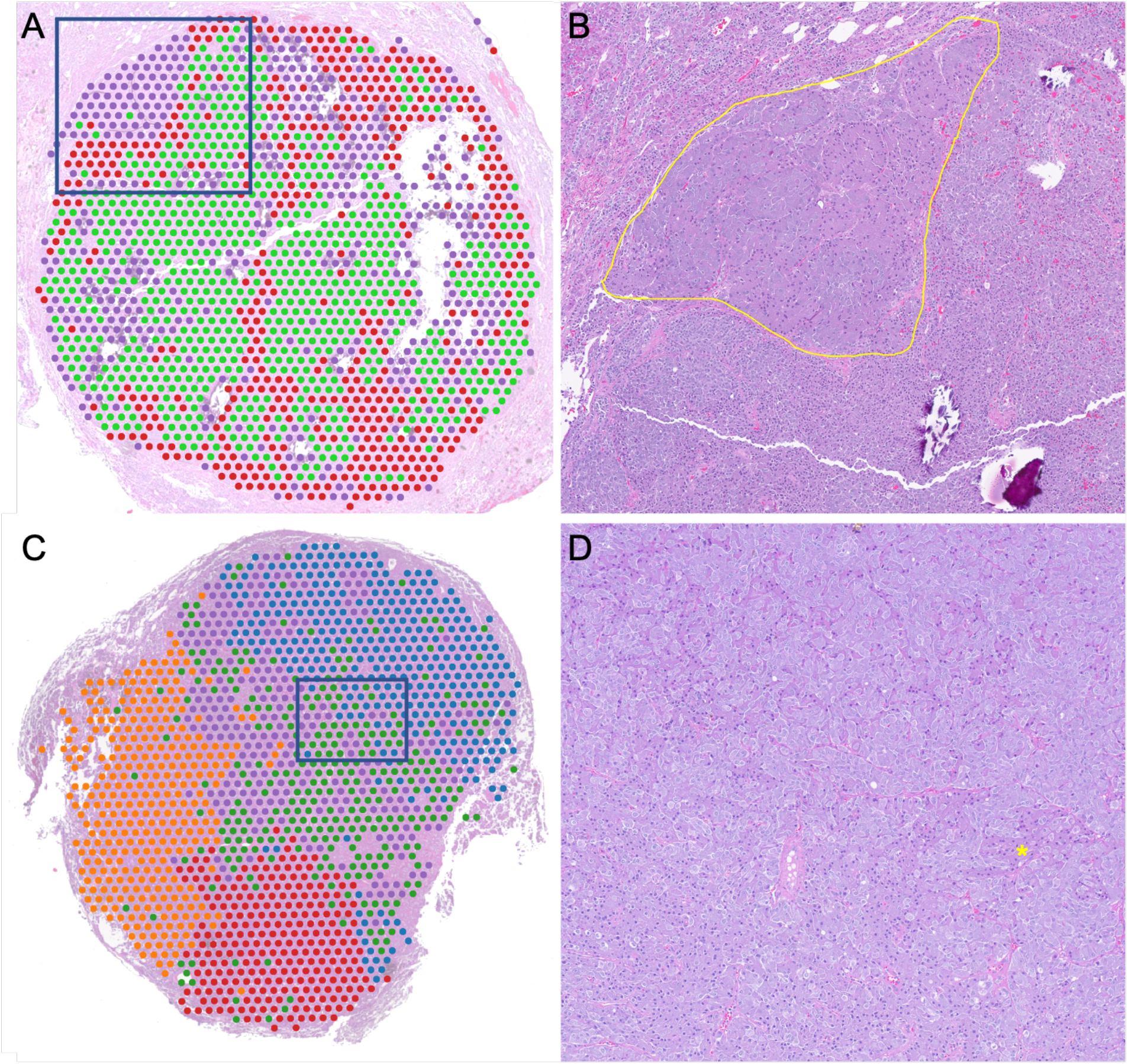
Intratumor cell heterogeneity. (A, C) In situ intratumor cell cluster annotations (each cluster shown with different color). (B, D) H&E staining of the marked areas at the intersection of different intratumor cell clusters showing differences in cell characteristics. (B) Cells inside the yellow marked area corresponding to the purple cluster in (A) show different architectural organization and are morphologically larger than surrounding tumor cells. (D) Green spots in (C) correspond to a cluster of cells enriched in a cell population with more eosinophilic cytoplasm (representative cell population highlighted with yellow asterisk).

Following intratumoral gene expression-based clustering, an expert pathologist reviewed the H&E-stained sections for apparent morphologic differences between the intratumoral clusters. In some cases, such differences were identified; notably, none of them were included in the postoperative pathology report for these tumors. For example, in tumor 1D the cluster annotated with purple color corresponded to a different architectural organization of cells that were of larger size compared to the surrounding tumor (Fig. 4A, 4B). Further, in other tumors, subtle differences in cell morphology were noted upon close review of the H&E staining of populations in different clusters (Fig. 4C, 4D).

### Intratumor clustering - Gene profiles

Review of expression profiles of the intratumor clusters revealed that in certain cases, intratumor transcriptional clustering corresponded to variability in relative expression of the marker genes of corticotroph cells, indicating transcriptional heterogeneity and suggesting differential secretory capacity (Fig. 5B). There was no apparent correlation between the presence of this intratumoral functional heterogeneity and the circulating ACTH or urinary free cortisol levels of the patient.

**Fig. 5.**
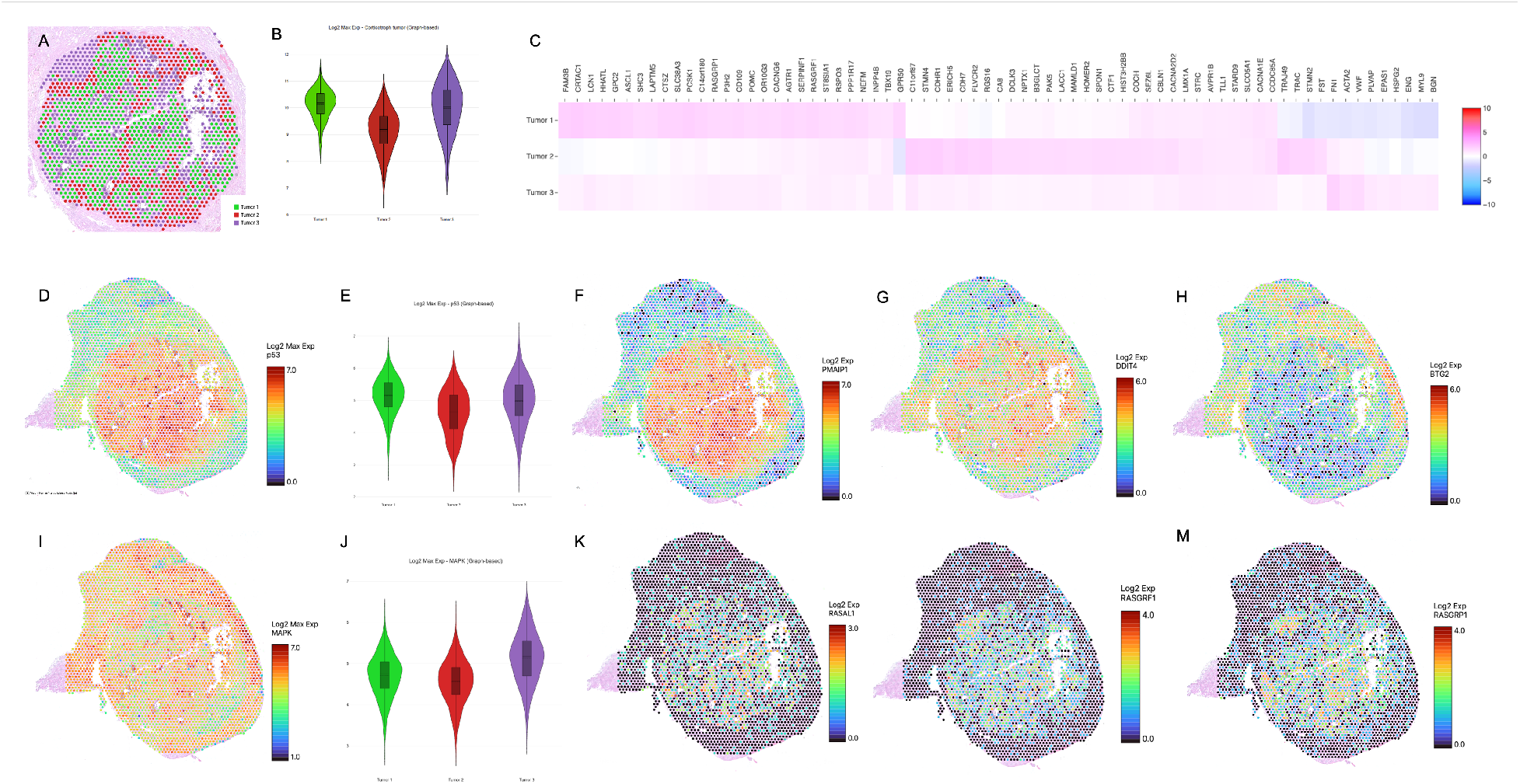
Expression profile of intratumor clusters. (A) In situ intratumor cell cluster annotations (each cluster shown with different color). (B) Violin plot of relative gene expression levels of corticotroph marker genes of the tumor clusters. (C) Heatmap of the top-ranked differentially expressed genes between the three tumor clusters. (D) In situ annotation and (E) violin plot of the relative expression of selected genes involved in p53 pathway. (F-H) In situ annotation of expression of select genes involved in the p53 pathway. (I) In situ annotation and (J) violin plot of the relative expression of selected genes involved in MAPK pathway. (K-M) In situ annotation of expression of select genes involved in the MAPK pathway.

We further reviewed the differentially expressed genes of each cluster and performed pathway analysis to identify genes or pathways of interest. Based on this analysis and taking into account the literature of known pathogenetic mechanisms of PitNETs, we focused on two pathways, the p53 and the mitogen-activated protein kinase (MAPK) and reviewed them more thoroughly on one sample (1D) to understand their in situ expression patterns.

Genes involved in p53 pathway showed differential expression among the tumor clusters but also diffusely within the tumor tissue (Figure 5D, 5E). Among the genes involved in p53 pathway that were more markedly differentially expressed, we noted differences in *PMAIAP1* (upregulated compared to normal gland, highest log2 fold change (Log2FC): 1.4, adjusted *P (AdjP)* < 0.001), *DDIT4* (upregulated compared to normal gland, highest Log2FC: 0.6, *AdjP*: 0.07), and *BTG2* (downregulated compared to normal gland, highest Log2FC: −1.3, *AdjP* < 0.001) (Figure 5F-H).

On review of components of the MAPK signaling pathway, it was evident that although normal pituitary gland showed higher overall expression of the cumulative expression of marker genes of MAPK pathway, different genes contributed to the expression profile in the tumor versus the normal surrounding tissue. Some of these distinct genes included *RASAL1* (Log2FC: 1.4, *AdjP* < 0.001), *RASGRF1* (Log2FC: 1.6, *AdjP* < 0.001), and *RASGRP1* (Log2FC: 1.5, *AdjP* < 0.001) which showed upregulation in the tumor with heterogeneous expression across the tumor tissue (Figure 5I-M).

## Discussion

ST analysis has emerged as a powerful tool for expression profiling of tumors with the additional benefit of providing information on the in situ organization of the cells in the context of their surrounding structures and microenvironment. We herein describe the ST analysis of corticotroph tumors that demonstrates unique information on intratumor heterogeneity.

PitNETs are considered to be predominantly monoclonal lesions, although data suggest that they may acquire overtime genomic changes that contribute to their behavior.(14,15) Cui *et al* studied intratumor heterogeneity using scRNA-seq.(9) Although the investigators identified small clusters of cells with differential expression in plurihormonal tumors and in one case of CD, these clusters did not reveal specific gene pathways and some markers were not confirmed in immunohistochemistry.(9) Asuzu *et al* studied corticotroph PitNETs and compared central versus marginal tumor samples identifying a list of differentially expressed genes.(8) We present here evidence that there is intratumor heterogeneity of the expression profile of tumor cells. The intratumor heterogeneity is often not based on central vs peripheral areas but rather in clusters or diffuse cell populations within the tumor.

It is possible that some of the differences we noted are due to intratumor hemorrhage and/or necrosis (subclinical apoplexy). These events are not uncommon in PitNETs and may lead to changes in their biochemical presentation.(16) This could also contribute to the differences in gene expression between clusters. Indeed, we noted that certain clusters showed enrichment in genes involved in mitochondria oxidation, such as the clusters involving the cells with morphological differences in Fig 4C-4D, which is a hallmark of necrosis often noted in tumors. Further artifact from tissue sectioning that leads to fragmentation could also contribute to differences in gene expression; this is sometimes difficult to avoid when handling small tumors. However, it is notable that the expression of hallmark genes is conserved, providing a validation that the overall transcriptional identity is retained.

Given the known implicated pathways in pituitary tumorigenesis, we focused on two of them and reviewed their expression profiles within the same tumor. The p53 pathway is critical for cell cycle regulation. Upregulation of the p53 pathway is often a response to cell stress and aims to control cell proliferation via induction of apoptosis, arrest of cell cycle and enhancement of DNA damage repair mechanism.(17) P53 pathway defects are found in more than half of human cancers and have been previously implicated in tumorigenesis of PitNETs.(4,17) Increased staining of p53 also correlates with more aggressive behavior in PitNETs.(18) Of the p53 pathway genes with high differential expression, *PMAIP1* has been recently reported by Asuzu *et al* as a component of the apoptosis regulation in corticotroph PitNETs.(8) Additionally, we noted that upregulation of *DDIT4* gene follows a similar pattern. *DDIT4* codes for the DNA damage inducible transcript 4 (DDIT4) protein which interacts with p53 protein but also plays an important role in PI3K-Akt-mTOR pathway and the Ras-signaling. High expression of DDIT4 in certain cancers is associated with poorer outcome and may be used as prognostic biomarker.(19) Finally, BTG2 is a pro-apoptotic factor regulated by TP53. Downregulation of *BTG2* is a hallmark of several human cancers suggesting an important role on cell proliferation and tumor growth.(20)

Furthermore, the MAPK pathway is considered pivotal for the response to a variety of growth factors or other stimuli leading to increased cell growth, and it has been previously implicated in PitNETs, mostly of the somatotroph or lactotroph lineage.(21) We here highlight specific components of the pathway that show differential distribution within the tumor and regulate Ras oncogene activity, such as the Ras protein activator like 1 (*RASAL1*), the Ras protein specific guanine nucleotide releasing factor 1 (*RASGRF1*), and the mediator Ras guanyl-releasing protein 1 (*RASGRP1)*.(22,23) This pathway is of particular interest since it has been targeted for therapeutic approaches in several cancers and specifically in PitNETs (24). Medications inhibiting the pathway, such as vemurafenib (selective BRAF V600E inhibitor), trametinib (selective MEK1/2 inhibitor) and gefitinib (selective EGFR inhibitor) are available. (25) Thus, it is important to note the intratumor heterogeneity of the pathway expression that could have implications on therapeutic response.

This study provides also the first validation of the feasibility of ST technique in pituitary tumors. We demonstrate that transcriptomic information deriving with this method can identify accurately the tumor location similar to standard staining as well as correlate with other critical markers of the tumor analysis (reticulin framework breakdown, Ki-67 index). Although time- and cost-effectiveness remain favorable for immunostaining at this time, it provides initial validation of the technique and confirmation that the gene expression profile of tumor cells is retained and correctly annotated using ST. As corticotroph PitNETs are often small with a median diameter of 5mm,(26) in the event that there is limited tissue available for research purposes, this technique could provide a genome-wide expression profile without the need for multiple stains or the need to select only certain markers of interest.

One of the limitations of this method is that it does not provide single cell resolution of the transcriptional profile and we noted that in certain cases areas of tumor and normal pituitary gland cells were pooled in one cluster. The accurate cell deconvolution of these areas required analysis of specific marker gene expression. Incorporating genetic information from scRNA-seq can help in correct annotation of these cell populations. Additionally, we do not have DNA genetic analysis of the tumors and further correlation of the identified transcriptional profile, the intratumor heterogeneity and the genetic mechanism leading to tumorigenesis would be important to evaluate.

To conclude, our results support that ST analysis of PitNETs unveils another dimension of the tumor development and progression. ST of tumors reveals various cell clusters inside the same tumor which may suggest variable cell differentiation pathways and may have implications for disease prognosis and therapeutic responses.

## Methods

### Patients

Patients enrolled under study 97CH0076 (Principal Investigator: Dr. Christina Tatsi, Clinicaltrials.gov Identifier NCT00001595) were reviewed. Seven patients with diagnosis of CD who underwent surgical resection of a corticotroph tumor within the previous 2 years, and for whom there was available surgical material were included in the study. Diagnosis of CD was based on the Endocrine society criteria and adjusted for the pediatric population as previously described.(27,28) Patients underwent surgery at the NIH CC and histologic reports of the resected material confirmed in all cases the presence of a corticotroph PitNET. Remission was defined as postoperative cortisol nadir <2 mcg/dL.

Written informed consent was obtained from parents and assent from patients if developmentally appropriate. All study procedures were approved by the *Eunice Kennedy Shriver* National Institute of Child Health and Human Development (NICHD) Institutional Review Board (IRB). Sample ID’s and codes were randomly generated and deidentified for downstream processing and analysis in this study.

### Immunostaining assays

Routine staining was performed on FFPE sections per standard laboratory techniques. H&E stains were performed on sections close to the one used for spatial transcriptomics, but other stains may have been on cuts further from the spatial transcriptomic slide. All immunohistochemistry procedures were performed in Ventana Ultra Autostainers (Roche Diagnostics, Indianapolis, IN). Ki-67 index was calculated in HALO Link application.

#### Spatial RNA Sequencing (spRNA-seq, 10x Genomics, Visium)

FFPE samples were selected based on their size, time of storage, and if available the previous report of increased Ki-67 index as a marked of increased cell proliferation. FFPE samples were cut and sectioned directly onto the 10x Genomics barcoded Visium slides (10X genomics part# 1000338). Two Visium slides were used, each containing four capture areas that were 6.5 x 6.5mm^2^. Every capture area consisted of approximately 5,000 gene expression spots with primers for sequencing and identification in downstream analyses. Seven of the eight panels contained tumor samples used for this study. Six of the seven total capture areas contained a single pituitary tumor. The remaining capture area had two tissues: a pituitary tumor and a small piece of normal gland used as a control.

All steps from 10X Genomics’ FFPE Visium Spatial workflow-specific protocols were followed. In short, after the slices were placed on the slides, they were then deparaffinized, stained with Hematoxylin & Eosin (H&E), imaged, and decrosslinked according to the Demonstrated Protocol (10x Genomics-protocol CG000409 Rev B). Samples were processed using the Visium Spatial Gene Expression Reagent Kits for FFPE (10x Genomics – protocol CG000407 Rev D). A human whole transcriptome probe panel was used to hybridize probes to their complementary RNA on the tissue sections. The probe pairs were ligated together and released from the tissue through RNAse treatment and permeabilization before getting captured onto the Visium Slide. The ligation products were extended and consisted of partial read 1, 10X Genomics spatial barcode, unique molecular identifier, Poly A, probe insert, and partial read 2 sequences. The ligated probe products underwent a sample index PCR to add P5, i5, i7, and P7 sequences needed for the Illumina platform. Sequencing was performed using the Illumina NovaSeq 6000 at a minimum of 25,000 read pairs per tissue-covered spot on the capture area. The sequencing saturation was above 40% for all samples with a median of 91,447 reads and 6,494 expressed genes per spot of tissue.

All differential spatial sequencing analyses were conducted using the Cell Ranger analysis pipeline. Principal component analysis (PCA) was run on the feature barcode matrices and clustering was reported based on the minimum number of gene dimensions. Results, including specific gene or gene list expression profiles and uniform manifold approximation and projection (UMAP) were reviewed at the 10X Genomics Loupe Browser. Lists of select marker genes used in certain analyses (marker genes of corticotroph cells, non-corticotroph pituitary lineages, collagen and fibroblasts, and genes of p53 and MAPK pathway as listed in Wiki pathway database) are provided in supplementary material. Gene pathway analysis was performed on differentially expressed genes for each slide in R using the *clusterprofiler* package for gene set enrichment analysis. Wiki, Hallmark and KEGG pathways were reviewed to identify enriched pathways within each cluster and identify possible pathways of interests involved in these processes.

## Data availability

The data that support the findings of this study are available either in Supplementary data or from the corresponding author upon reasonable request.

## Author contributions

C.T. and F.R.F. designed the study; F.R.F., J.O.P., and C.P. performed experiments; C.T., C.S.F., F.R.F, and J.O.P. analyzed the data; C.T. and P.C. provided clinical care for the patients; C.T. wrote the manuscript with input from all authors.

## Supplementary material

**Supp. Fig. 1.**
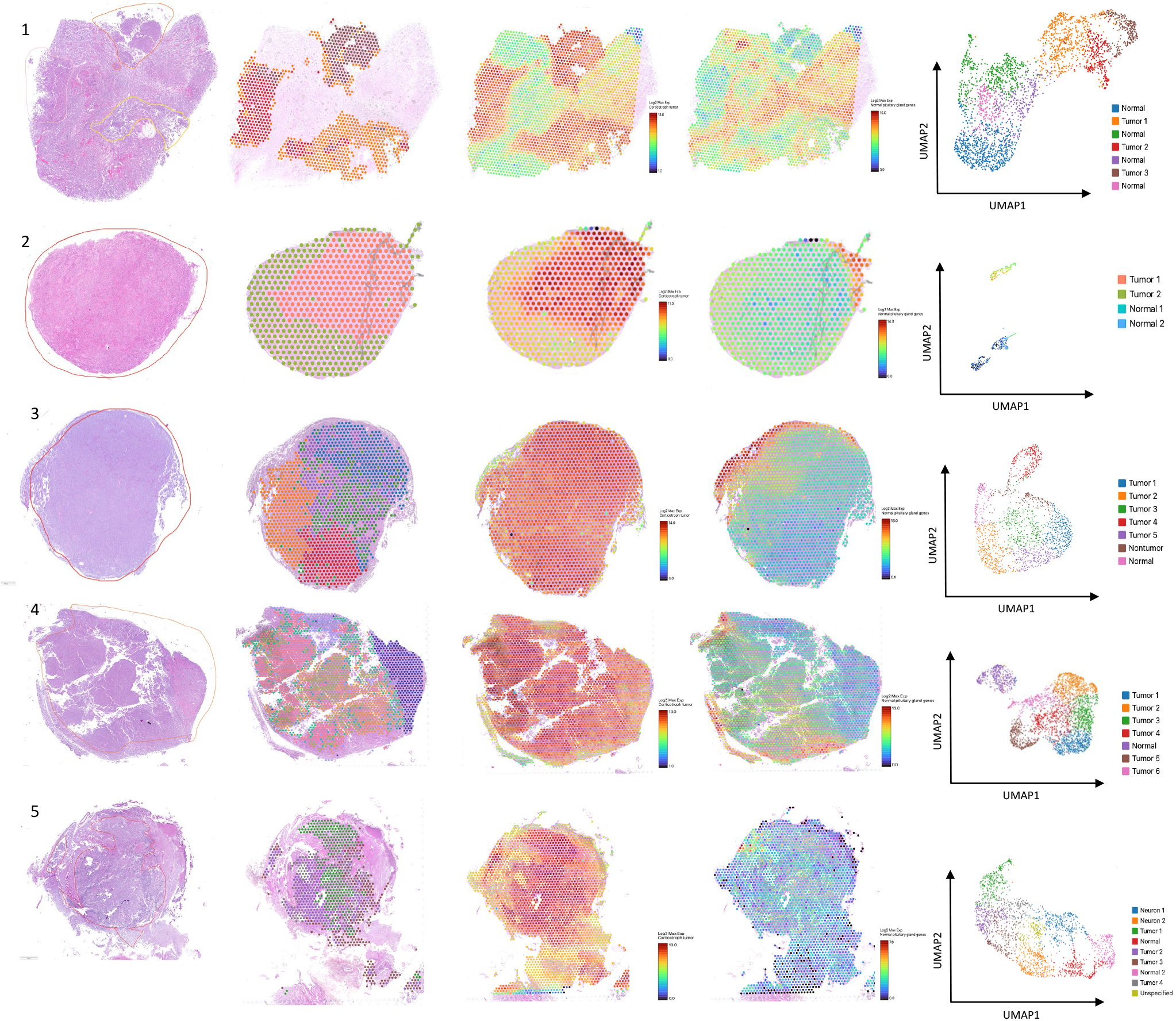
Images of pituitary tumors with pathologist’s annotation of tumor on H&E-stained slides (first column), cells clusters annotated as tumor (second column), gene expression profile of genes of corticotroph tumors (third column), gene expression profile of genes of normal pituitary gland (fourth column), and UMAP graph of the clusters annotated as tumor or normal gland (fifth column). Each row corresponds to the same tumor (Row 1: Patient 1A; Row 2: Patient 1B; Row 3: Patient 2B; Row 4: Patient 2C; Row 5: Patient 2D)

**Suppl. Dataset 1**. Gene lists used for analysis of expression profiles.

